# Detection of Drug Effect Signals Associated with Adverse Pregnancy Outcomes Using Propensity Score Matching at Scale

**DOI:** 10.1101/2024.03.21.24304579

**Authors:** Yeon Mi Hwang, Samantha N. Piekos, Qi Wei, Nathan D. Price, Leroy Hood, Jennifer J. Hadlock

## Abstract

**Objective:** We applied propensity score matching method at scale on patient records to confirm signals of known drug effects on preterm birth and detect previously unidentified potential drug effects.

**Materials and Methods:** This was a retrospective study on women who had continuity of care at Providence St. Joseph Health (PSJH) both before and after pregnancy and delivered live births between 2013/01/01 and 2022/12/31 (n=365,075). Our exposures of interest were all outpatient medications prescribed during pregnancy. We limited our analyses to medication that met the minimal sample size (n=600). The primary outcome of interest was preterm birth. Secondary outcomes of interest were small for gestational age and low birth weight. We used propensity score matching at scale to evaluate the risk of these adverse pregnancy outcomes associated with drug exposure after adjusting for demographics, pregnancy characteristics, and comorbidities.

**Results:** The total medication prescription rate increased from 58.5% to 75.3% (*P*<0.0001) from 2013 to 2022. The prevalence rate of preterm birth was 7.7%. 175 out of 1329 prenatally prescribed outpatient medications met the minimum sample size. We identified 58 medications statistically significantly associated with the risk of preterm birth (P≤0.1; decreased: 12, increased: 46).

**Discussion:** We narrowed down from 1329 medications to 58 medications that showed statistically significant association with the risk of preterm birth even after addressing numerous covariates through propensity score matching.

**Conclusion:** This data-driven approach demonstrated that multiple testable hypotheses in pregnancy pharmacology can be prioritized at scale, laying the foundation for application in other pregnancy outcomes.

## BACKGROUND AND SIGNIFICANCE

Pharmaceutical companies primarily rely on pre-marketing randomized clinical trials to prevent and assess adverse drug reactions (ADRs). Despite the effort, studies conducted on inpatient populations estimated a serious ADRs incidence rate of 6.7% (N ≥ 2,216,000) with a fatality rate of 0.32% (N ≥ 106,000), placing ADRs as the fourth leading cause of morbidity and mortality in the United States (US) health care systems.[1,2] The incidence rate of ADRs in outpatients is harder to estimate, with studies suggesting rates ranging from 3% to 38%.[3–8] Estimated incidence rate of ADRs in both inpatient and outpatient demonstrates that unintended drug response is common and expected.

Pre-marketing random clinical trials rarely include pregnant women unless the product targets pregnant women.[9] Consequently, drug efficacy, safety, and dosages are determined based on data from men and non-pregnant women. While pregnant women are the most underrepresented population in clinical trials, they can experience some of the most complex medical situations. During pregnancy, women undergo marked physiological changes that significantly alter the pharmacokinetics and pharmacodynamics of drugs.[10] Therefore, current knowledge in pharmacology should not be directly applied to pregnant women, as inadequate information on the pharmacology of pregnancy exposes them to a high likelihood of experiencing unintended drug responses.

Despite the limited availability of safety information regarding medication use during pregnancy, many pregnant women continue to use medications. Overall, 93.9% of pregnant women take at least one medication (over-the-counter or prescribed) and typically use an average of 4.2 during pregnancy.[11] Usage of prescribed medication by pregnant women varies globally, ranging from 23% to 96%, with the US in 2008 reporting a usage rate of 49% among pregnant women.[11] Given the prevalent use of medication among pregnant women and the challenges associated with conducting prospective clinical trials on this population, leveraging real-world data has emerged as a promising supplemental approach to investigate the effects of drugs during pregnancy. Electronic health records (EHRs) are particularly suitable candidates among these real-world data sources. EHRs contain rich and comprehensive information about patients’ longitudinal health profiles, potential confounding factors, and prescription history. Active research on developing novel methodologies for not only ADRs[12,13] but also for drug repositioning[14] and drug-drug interactions[15,16] is ongoing.

However, despite these advancements in data-driven healthcare research, the field of pregnancy research has been slower in adopting these novel methodologies. In summary, there is a pressing need to establish a foundational framework for systematically investigating drug responses during pregnancy at scale using real-world data. Such an effort is crucial, as it can lead to the generation of testable hypotheses related to drug effects on pregnancy outcomes, both positive and negative. Furthermore, uncovering drug responses that do not pose risks to adverse pregnancy outcomes can provide valuable insights into drug safety during pregnancy. Here, we selected preterm birth (PTB) as our primary outcome of interest. PTB, defined as birth occurring before 37 weeks of gestation, significantly contributes to perinatal morbidity and mortality in developed countries. PTB accounts for 75% of perinatal mortality cases and over half of long-term morbidity.[17]

## OBJECTIVE

We employed a large-scale propensity score matching approach on patient records to expedite the generation and prioritization of testable hypotheses related to the risk of PTB. We hypothesized there exist not yet characterized pharmacological signals with medication and risk of PTB. Beyond hypothesis generation, we investigated a few detected drug effect signals using traditional pharmacoepidemiology methods.

## MATERIALS AND METHODS

### Study design, setting, and participants

Providence St. Joseph Health (PSJH) is an integrated US community healthcare system that provides care in urban and rural settings across seven states: Alaska, California, Montana, Oregon, New Mexico, Texas, and Washington. We used PSJH pregnant patient records who delivered live infants from January 1, 2013, through December 31, 2022 (n=543,408). Figure S1 describes the cohort selection. We excluded multiple pregnancies and deliveries with gestational age (GA) of less than 20 weeks (n=516,881). GA was limited to 20 weeks or greater because ascertainment bias is particularly high for EHR data earlier in pregnancy. We limited our analyses to pregnant patients aged between 18 and 45 years of age (n=510,488), who had continuity of care at PSJH before and after pregnancy. We included patients who had continuity of care: at least one encounter 180 days before the start of pregnancy (last menstrual period, LMP) to the time of delivery (n=365,075). This was done to partially address surveillance bias.

All procedures were reviewed and approved by the Institutional Review Board at the PSJH through expedited review on 11-04-2020 (study number STUDY2020000196). Consent was waived because disclosure of protected health information for the study involved no more than minimal risk to the privacy of individuals.

### Variables

### Exposures

We mapped all prescription records during pregnancy to RxNorm code based on ingredients. We split the cohort into exposed and unexposed groups for individual medication ingredients. Women with medication orders that overlapped with at least one day of pregnancy were considered exposed. We excluded medications that did not reach a minimum sample size of the exposed, which was 600. This minimum sample size was calculated using Epitools,[18] with the following parameters: PTB prevalence rate of the PSJH maternity cohort (7.7%), assumed relative risk (1.55), desired level of confidence (0.9), and desired power for the detection of significant difference (0.8). The calculated minimum sample size was 582, but we rounded it to 600.

### Outcomes

The primary outcome of interest was PTB, defined as gestational age at birth (GA; GA<37 weeks). Secondary outcomes were low birth weight (LBW; birth weight <2,500g) and small for gestational age (SGA; birth weight < 10th percentile of based on gestational age).

### Covariates

We extracted maternal, pre-pregnancy, and prenatal characteristics and comorbidities information from EHR data. Pregnancy and maternal characteristics were collected during prenatal care or at time of delivery. These included parity, preterm history, delivery year, fetal sex, age at LMP, race, ethnicity, insurance status, pregravid body mass index, smoking, and use of alcohol and illegal drugs (Table S1).

We conducted a parallel analysis with three different sets of covariates. First, we conducted propensity score matching with the covariates without comorbidities. Second, we addressed pre-pregnancy comorbidities based on the obstetric comorbidity index.[19] Selected comorbidities were renal diseases, chronic lung diseases, diabetes, leukemia, pneumonia, sepsis, cardiovascular diseases, sickle cell diseases, anemia, cystic fibrosis, and asthma(Table S2). A similar practice was done in an at-scale study conducted by Sentinel System, one of the US Food and Drug Administration (FDA) efforts in surveillance medical products.[20] We excluded comorbidities specific to the prenatal period, such as gestational diabetes; the obstetric comorbidity index is designed to assess the mortality risk at delivery. Third, we selected the fifteen most common comorbidities before and during the pregnancy. We acknowledge prenatal comorbidities do not satisfy the covariate definition. However, this study aims to explore the usefulness of EHRs and generate hypotheses. To do so, we employed an exploratory approach beyond the conventional one.

### Analysis

#### Descriptive statistics

We described the source population on maternal characteristics, outcomes, and covariates. The descriptive statistics are presented in Table S3. We characterized the prescription rate within the PSJH pregnant population in Figure 1. We used the chi-square test and linear regression to evaluate the difference in prescription rate across categorical variables and continuous variables. Age distribution of this source population is described in Figure 2. Prescription patterns from 2013 to 2022 based on their ingredient and ATC classification categories are displayed in Figure 3.

**Figure 1.**
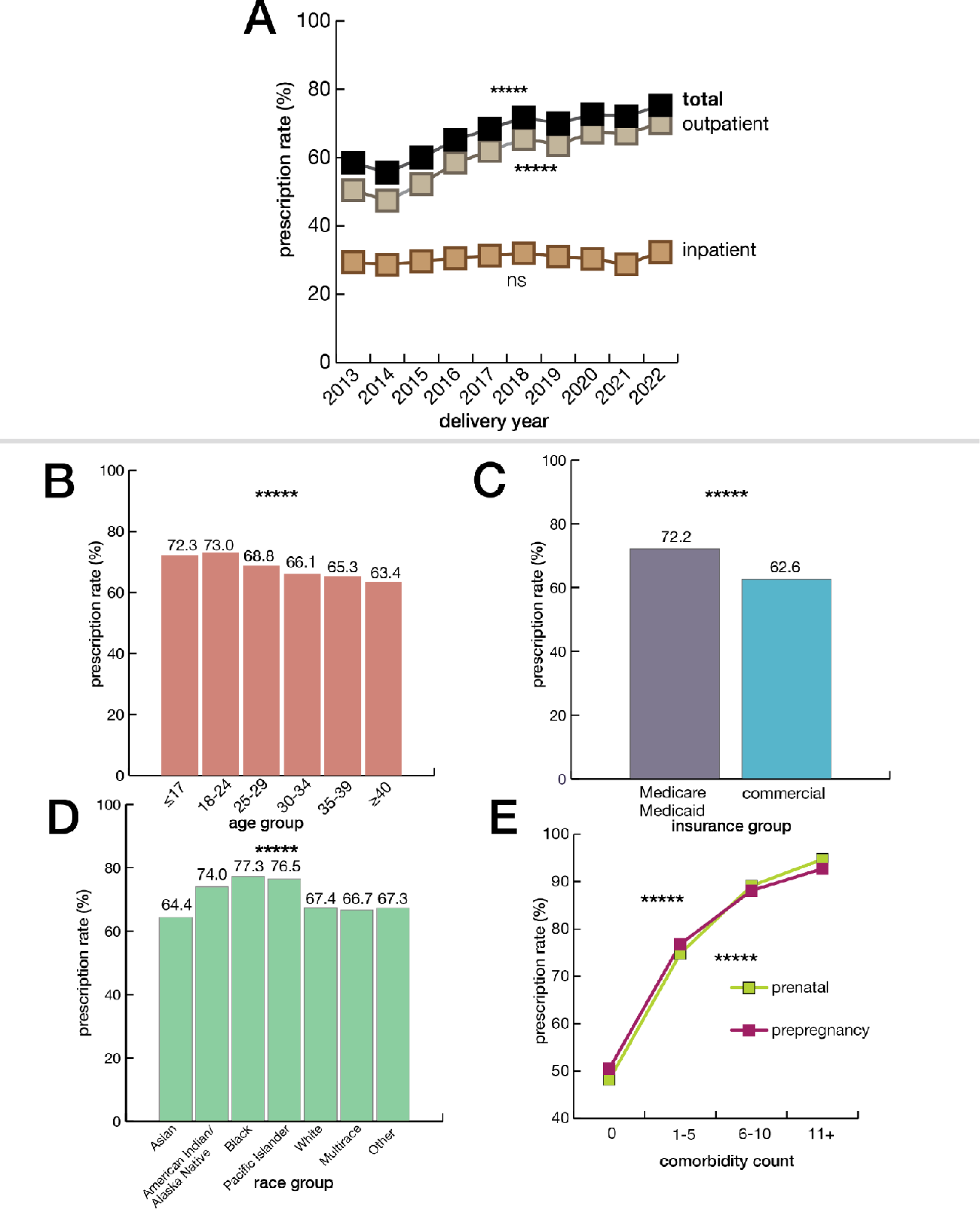
Overall prescription rate of PSJH pregnant population. (A) plot shows the increase in total prescription rate from 2013 to 2022. The total medication prescription rate increased from 58.5% to 75.3% from 2013 to 2022 (P<0.0001). The inpatient prescription rate increased from 29.3% to 32.4% (P=0.2). In contrast, outpatient medication prescriptions increased from 50.5% to 70.1% (P<0.0001). We evaluated the increase in prescription rate using linear regression. (B) plot shows the total prescription rate across age groups (P<0.0001). We evaluated the decrease in prescription rate across ages using linear regression. (C) plot shows the difference in prescription rates between insurance groups (P<0.0001). We evaluated the difference in prescription rate across categorical variables using the chi-square test. (C) plot shows the difference in prescription rate across race groups (P<0.0001). We evaluated the difference in prescription rate across categorical variables using the chi-square test. (D) plot shows the increase in prescription rate based on comorbidity count. The increase in prescription rate across comorbidity count using linear regression.

**Figure 2.**
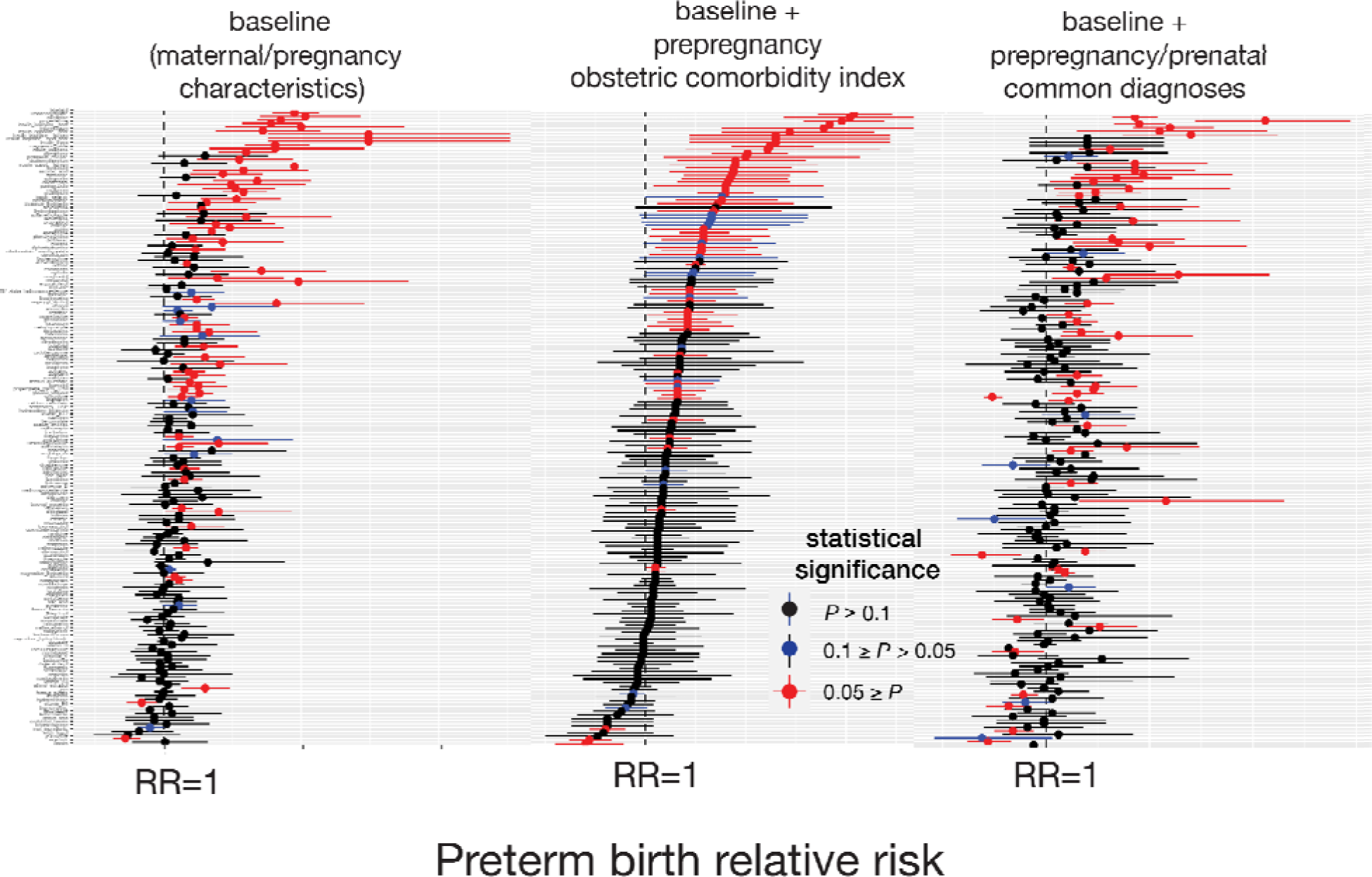
Forest plots of association between medication and risk of PTB. Left plot shows the forest plot of baseline analysis that adjusted maternal and pregnancy characteristics. The center plot shows a forest plot of analysis that adjusted for maternal and pregnancy characteristics and pre-pregnancy comorbidities from the obstetric comorbidity index. The right plot is a forest plot of analysis that adjusted for maternal and pregnancy characteristics and prenatal/pre-pregnancy common comorbidities. The Y axis is the list of medications that met the minimum sample size in descending order of RR of analysis in the center plot. This figure is summarized in Table 1. RR, confidence interval, and p-values are reported in supplementary data.

**Figure 3.**
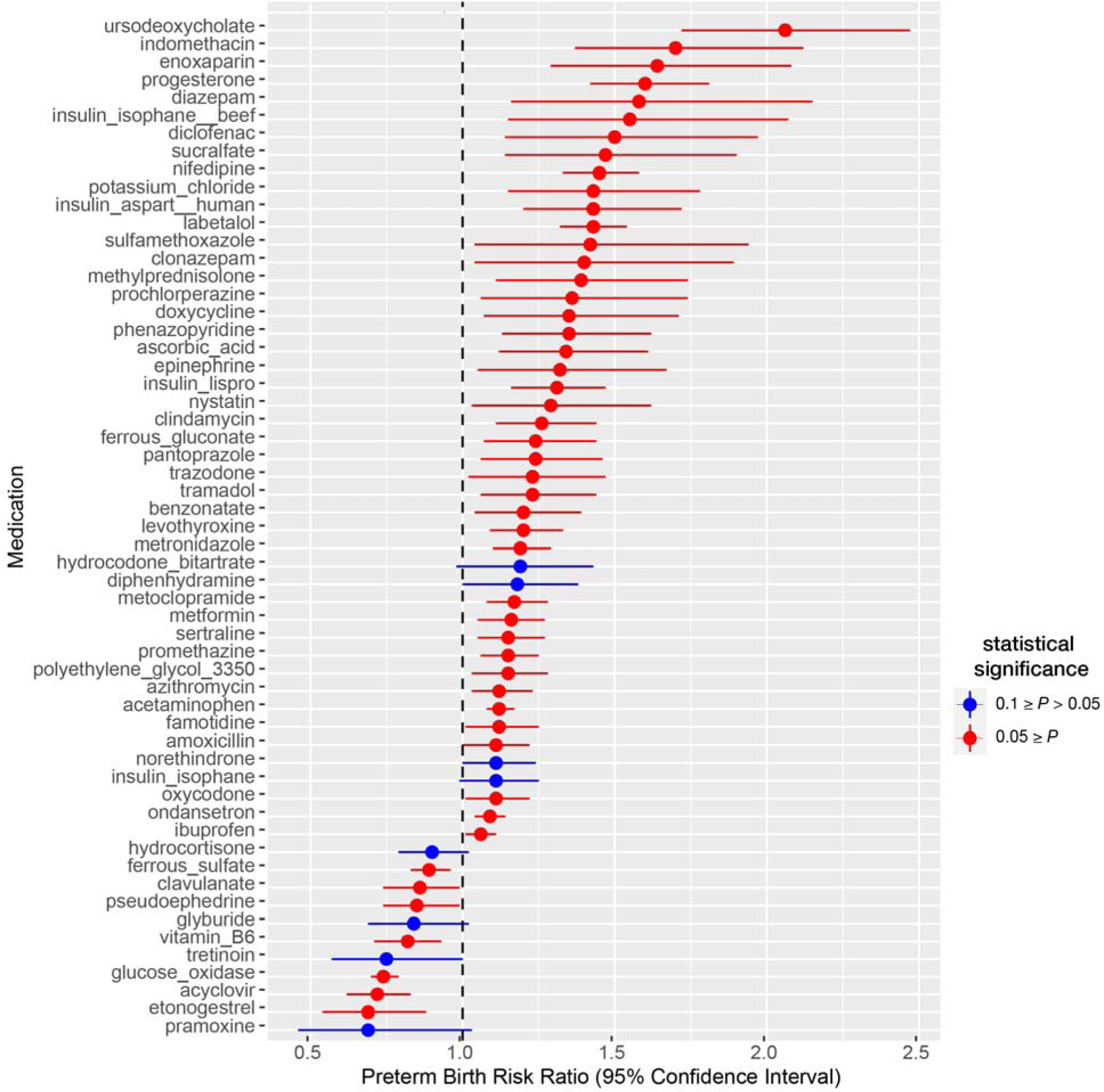
Forest plot of statistically significant association with risk of PTB.

**Table 1.**
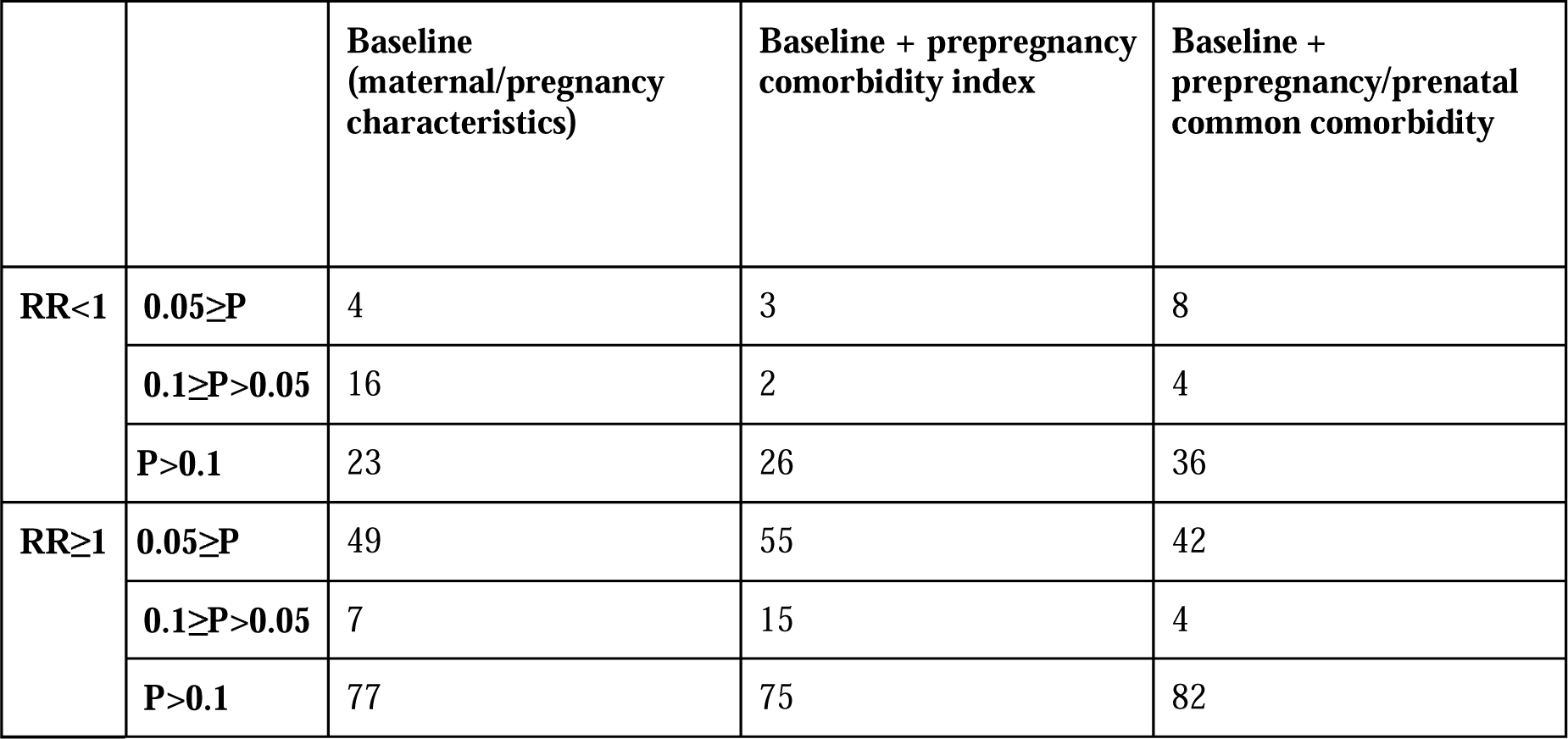
Summary of associations based on statistical significance and relative risk.

#### Propensity score matching

We calculated the risk ratio of PTB, LBW, and SGA for individual outpatient medications that reached the minimum sample size. For each medication, the unexposed group was matched to the exposed group on the covariates. We used propensity score matching to account for covariates associated with adverse pregnancy outcomes. Compared to other propensity score methods and covariate adjustment methods, propensity score matching provided exceptional covariate balance across most circumstances.[21] An unsupervised learning model with k-nearest neighbors (k=1), as recommended by a prior study,[22] was used to match with replacement by the propensity logit metric. We evaluated the covariate balance using an average standardized mean difference. We excluded medication ingredients with an average standardized mean difference below 0.2. We categorized medications with statistically significant associations based on their indication in three categories: preterm labor(PTL) or PTB, PTB risk factors, and infection(Table S4). Here, we considered association with a P value below 0.1 statistically significant. This is not a conventional practice in hypothesis-testing studies, but our study is hypothesis-generating. We are suggesting potential hypotheses for researchers to investigate further.

#### Validation

We selected sertraline, acyclovir, and ferrous sulfate for further investigation. They had relatively large exposure groups and were statistically significant in an analysis adjusted for pre-pregnancy/prenatal common diagnoses. Details of the method are described in the supplemental methods.

#### Sertraline

Sertraline is a Selective Serotonin Reuptake Inhibitor (SSRI) antidepressant. Depression is a treatable disease and a risk factor for PTB.[23] We limited our analytic population to patients who had any depression diagnosis before pregnancy(Table S2). We evaluated the risk of PTB in patients exposed to sertraline among patients who had depression onset before the pregnancy. Additionally, we assessed the likelihood of delivering preterm in patients exposed to SSRI within the same analytic population that we used to evaluate the risk of PTB in those exposed to sertraline.

#### Acyclovir

Acyclovir is a treatment for herpes virus infection, including shingles, chicken pox, and genital herpes. Genital herpes is a sexually transmitted disease, which is a risk factor for PTB. We determined the indication of treatment based on dosage.[24] According to the CDC treatment guideline,[25] acyclovir is recommended starting at GA 36 weeks to suppress the reactivation of genital herpes among pregnant women. Patients who adhered to this treatment guideline delivered after 36 weeks of gestation, potentially introducing selection bias and leading to a lowered risk of PTB. Initially, we characterized the number of patients who initiated their prescription at 36 weeks of gestation to assess the proportion of patients following this CDC treatment guideline. Subsequently, we examined the likelihood of PTB in patients exposed to acyclovir before 36 weeks of gestation. We replicated the analysis on a subsample of patients who had indications of genital herpes (Table S2). We then evaluated the risk of PTB among patients exposed to acyclovir or valacyclovir (oral prodrug of acyclovir) before 36 weeks of gestation.

#### Ferrous sulfate

Ferrous sulfate is a treatment for iron deficiency anemia, which is a risk factor for PTB. We assessed the impact of ferrous sulfate in the anemic group. The anemic group was determined based on the presence of iron-deficiency anemia diagnosis within 180 days before LMP to LMP (Table S2).

## RESULTS

### Descriptive statistics

We identified 365,074 patients with singleton pregnancies and continuity of care as our analytic population. This population was enriched with people who were aged 30-34 (32.7%), White or Caucasian (63.2%), non-Hispanic or Latino (77.2%), Medicaid/Medicare insured, living in metropolitan areas (84.2%), and delivered in 2022 (12.4%). Median maternal age increase from 30.3 to 31.5 (P<0.0001) from 2013 to 2022. The proportion of women aged 35 or older increased from 20.8% to 27.0% from 2013 to 2022(Figure S2). The mean gestational age at delivery was 275.0 days. The average prevalence rates of PTB, SGA, and LBW were 7.7%, 12.1%, and 5.4%.(Table S3)

The total medication prescription rate increased from 58.5% to 75.3% from 2013 to 2022 (P<0.0001). The inpatient prescription rate slightly increased from 29.3% to 32.4% (P=0.2) In contrast, outpatient medication prescriptions increased from 50.5% to 70.1% (P<0.0001) (Figure 1). The maternal age group of 18-24 had the highest prescription rate of 73.0%. Mothers aged 40 or older had the lowest prescription rate reporting 63.4%(P<0.0001). The Medicare/Medicaid insurance group had a higher prescription rate reporting 72.2%, than the commercial insurance group (62.6%; P<0.0001). Amongst the race group, pregnant women who reported Black or African American race had the highest prescription rate of 77.3%, and Asian had the lowest, reporting 64.4%(P<0.0001). We observed prescription rate increases as the number of comorbidities increased. This trend was similar for both pre-pregnancy and prenatal comorbidities. Approximately half of the pregnant people with no pre-pregnancy/prenatal problem diagnosis had a prescription during pregnancy. Patients with eleven or more pre-pregnancy/prenatal problem diagnoses had a prescription rate higher than 90% (Figure 1).

### Propensity score matching

From the initial pool of 1329 medications, 175 prenatally prescribed medications met the minimum sample size. None of the medications had an effect size below 0.2 after matching all three analyses. When we adjusted for baseline characteristics, pregnancy and maternal characteristics, we identified a total of 76 (RR<1: 20, RR≥1:56) associations with a p-value below 0.1.(Table 1) The number of associations with statistical significance narrowed when additionally accounting for pre-pregnancy comorbidities in the obstetric comorbidity index. We observed 75 (RR<1: 5, RR≥1:70) medications associated with the risk of PTB with statistical significance. (Table 1) Finally, we identified 58 (RR<1: 12, RR≥1:46) medications associated with the risk of PTB in an analysis adjusted for common diagnoses during the pre-pregnancy and prenatal period (Figure 2, Figure 3, and Table 1). Statistically significant correlations were categorized into three categories based on their indication: PTL/PTB, risk factor of PTB, and infection.(Table S4) Forty-three medications had indications categorized into at least one category. Four medications fell into the category of PTL/PTB indication. Thirty-two medications had indications that were risk factors for PTB. Nine medications were prescribed in case of infections, including bacterial, fungal, and viral.

### Validation

#### Sertraline

There were 29,352 patients who had depression diagnosis before the pregnancy. Respectively, 3214 and 5910 patients were exposed to sertraline or any SSRI. They were 1.28 times [1.14, 1.45] and 1.16 times [1.05, 1.28] more likely to deliver preterm than patients without exposure.

#### Acyclovir

The majority of patients (58.8%; 4947 out of 8420) who had prenatal acyclovir exposure started their prescription at or after 36 weeks of gestation. Those exposed to acyclovir before 36 weeks of pregnancy had 1.77 times (1.77 [1.52, 2.07]) higher likelihood of delivering preterm compared to patients without prenatal acyclovir exposure. However, within the subsample of patients diagnosed with genital herpes, we did not observe an elevated risk of PTB (OR=1.19 [0.94, 1.50]). Additionally, there was no observed association between exposure to acyclovir and elevated risk of PTB when comparing individuals exposed to acyclovir and valacyclovir before 36 weeks of gestation (OR=0.86 [0.74, 1.00]).

#### Ferrous sulfate

There were 774 patients were diagnosed with iron deficiency anemia within a 180-day pre-pregnancy period. We observed 294 patients with a prescription for ferrous sulfate during pregnancy. Our analysis revealed no association between the prescription of ferrous sulfate and the risk of PTB (OR=0.85[0.48, 1.50])

## DISCUSSION

To our knowledge, this was the first study to use propensity score matching at scale on EHR to generate and prioritize testable hypotheses on drug effects associated with risk of PTB. We retrospectively assessed 365,074 people who were continuously enrolled in PSJH. The majority of women took prescribed medication during pregnancy. From an initial pool of 1762 medications, we narrowed down to 172 medications for hypothesis evaluation. Three of these detected signals were selected based on their relatively large exposure groups and statistical significance in an analysis adjusted for pre-pregnancy/prenatal common diagnoses. We evaluated the heightened likelihood of delivering preterm associated with exposure to sertraline and decreased chance related to exposures to acyclovir and ferrous sulfate.

We employed propensity score matching at scale on EHR and produced hypotheses for 172 medications. Among them, 57 of 172 mediations had statistically significant associations with the risk of PTB. There were a few prior studies with similar aims. Maric et al., 2019[26] assessed administrative claims data on 2,538,255 deliveries and identified 863 medications with statistically significant associations. Their number of signals, statistically significant association, far exceeds ours because their sample size was greater and did not eliminate medication that did not meet the minimum sample size. That study had only 5 medications with an odds ratio below 1, whereas we had 12. Another effort to establish a framework to detect drug effect signals in maternal-fetal medicine was conducted by the Sentinel working group. Sentinel initiative, led by the US Food and Drug Administration (FDA), has created novel methods to evaluate the safety of approved medical products, including medications, vaccines, and devices. They used propensity score matching tree-based scan statistics methods on Medicaid data to discover infant outcomes associated with prenatal cephalosporin exposure in the first trimester.[20] That study utilized a different approach as they focused on multiple outcomes and single exposures; our study assessed single outcomes and multiple exposures. Both prior studies utilized claims data, whereas we used EHR.

The majority of patients were prescribed medications during pregnancy. This finding corresponds to observations in earlier studies. According to Mitchell et al. 2011,[11] in the US, 48% of women were exposed to prescribed medication during pregnancy in 2008. A systemic review study conducted on peer-reviewed literature from 1989 to 2010 in developed countries reported that 27% to 93% of pregnant women used prescription drugs, depending on the country.[27] In our study, we observed an increase in prenatal prescription rate from 58.5% to 75.3% from 2013 to 2022. This rate is higher than the prescription rate reported in 2008. The discrepancy in the prescription rate for medication during pregnancy may be attributable to a gradual increase in usage. Mitchell et al. in 2011 described an incremental increase in the use of prescription medications by 60% from 1986 to 2008. We also observed a rise in the prescription rate from 2013 to 2022. As discussed in the introduction, the common use of medication during pregnancy underscores the necessity to promote pharmacology research in pregnant women and to leverage already generated real-world data to expand our understanding of the efficacy and safety of medications during pregnancy.

Surprisingly, the prescription rate decreased as the maternal age increased. We first assumed that the increased prescription rate over the study period was attributable to increasing maternal age based on observation from Mitchell et al. 2011.[11] Indeed, the median maternal age increased, and the proportion of women aged 30 or older gradually increased over our study period. However, the prescription rate did not correlate with the maternal age, contrary to our speculation. Women in the oldest age group, 40 or older, had the lowest prescription rate, whereas women aged 24 or younger had the highest. The major difference between our study and Mitchell et al. 2011[11] is the study period and population. Their observation was based on 5008 deliveries from 1997 to 2003 in the US. In contrast, our observation was relatively similar to that from a more recent study[28] on 2.3 million patients who delivered live births from 2000 to 2019. Their study reported that the most prevalent medication exposures (antibacterial agents, antiemetics, and contraceptives) during pregnancy had a prescription pattern across age groups like our study. The younger group, 24 or younger, had much higher prescription rates for these medications than those of the older group, 35 or older.

We employed traditional pharmacoepidemiology methods to evaluate the detected drug effect signals for sertraline, acyclovir, and ferrous sulfate. Specifically, we focused on assessing the negative association between sertraline/SSRI and the risk of PTB among patients who had an onset of depression before the pregnancy. We further validated this in a separate study.[29] We confirmed the correlation between exposure to sertraline/SSRI and the risk of PTB, and this correlation remained strong and significant through extensive sensitivity analyses. However, our study faced limitations in properly evaluating ferrous sulfate association with a lower risk of PTB due to the small sample size. Only 774 patients received a diagnosis of iron deficiency anemia within the 180-day pre-pregnancy period. Despite the small sample size of our study, a recent study reported that patients exposed to iron supplementation(ferrous sulfate, ferrous gluconate, ferrous fumarate, and ferrous glycinate) experienced reduced odds of preeclampsia and/or PTB.[30]

In contrast to sertraline and ferrous sulfate, the signal we observed for acyclovir was misleading. According to CDC treatment guidelines,[25] acyclovir is recommended for administration at 36 weeks of gestation for patients with genital herpes. This practice likely introduced selection bias, as the exposure group included patients who surpassed 36 weeks of gestation. In fact, 60% of patients were exposed to acyclovir at or after GA 36 weeks. When we restricted our exposure group to patients exposed to acyclovir before 36 weeks, the protective result associated with the risk of PTB disappeared. Interestingly, our result slightly differed from prior studies. In a previous study, exposure to valacyclovir, not to acyclovir, was associated with a lower risk of spontaneous PTB[26]. Investigations on sertraline and ferrous sulfate demonstrate the potential of our approach to produce and prioritize hypotheses to evaluate. However, misleading signals do exist. Thus, we must take a conservative stance and carefully verify detected drug effect signals with experiments designed to consider how the drug is used in a clinical context.

We identified 118 medications with no statistical significance. Restricting our analyses to medications that satisfy minimum sample size ensures that associations lacking statistical significance are not dismissed as meaningless. Considering that pregnant women are typically excluded from clinical medication trials despite their medication use, the absence of an association with the risk of PTB is a valuable finding supporting the potential drug safety associated with PTB. It underscores the need for similar studies in pregnancy pharmacology to be conducted and repeated on real-world data to gather more evidence on medication’s safety, risks, and benefits for pregnant women.

We had one of the largest sample sizes for hypothesis-generating retrospective EHR studies for pregnant women. While similar studies exist, they often rely on claims data.[20,26] Claims data may offer a larger sample size but EHR provides richer data on patient’s longitudinal health conditions, encompassing lab results, vital signs, and surveys.[31] Moreover, our study setting PSJH serves community hospitals and clinics in both rural and urban settings in seven western states in the US. This setting better reflects the general population than the academic hospital, which may have a greater proportion of high-risk pregnancies.

To ensure the integrity and reliability of our analyses, we implemented several measures to mitigate bias and ensure the robustness of our findings. We reduced the surveillance bias by restricting to continuously enrolled patients and leveraging propensity score matching. Furthermore, we mitigated the bias by matching patients in the treatment group to those in the control group with similar characteristics across covariates. Given that individuals exposed to medication may have more frequent doctor visits, ensuring comparability of patient health was crucial. Another noteworthy aspect of our approach was our commitment to evaluating all medications without introducing systemic bias. In research, there can be a tendency to focus on variables or hypotheses previously explored or considered more interesting. By conducting assessments on all medications that reached the minimum sample size, we aimed to prevent such biases from influencing our analysis, which contributed to the overall rigor of our study.

In this study, we deliberately did not correct for multiple testing errors because the primary objective of this study was to produce hypotheses, rather than to test them. Although multiple comparison can increase the likelihood of producing false positives, correcting this can increase the rate of false negatives. Furthermore, different methods for multiple testing correction can yield varying adjusted p-values. Instead of applying specific correction methods, we presented confidence intervals. This decision allows future researchers to use them for meta-analysis, as recommended by a prior study.[32] We underscore the need for cautious consideration of these associations and advocate for thorough evaluation through meticulously designed studies that reflect the characteristics of exposures of interest and their indications.

This study also has several limitations. By focusing on a study population of patients who received care within the healthcare system before pregnancy, we excluded transient patients admitted for delivery who were likely to lack prenatal information. This study population may be slightly biased toward lower-risk pregnancy cases, as some higher-risk pregnancy cases may have transitioned care to academic medical centers before delivery. Another limitation was the use of uniform sets of comorbidities for all medications. Although we conducted multiple analyses with several groups of comorbidities, it is essential to note that individual medications are prescribed for specific indications. Medications with fewer common indications may not be adequately represented in the covariate we investigated. In the future, we can address this limitation by applying several promising approaches. One such approach is high-dimensional propensity score matching.[33] High-dimensional propensity score matching offers a robust way to control for confounding variables in observational studies. Unlike traditional propensity score matching, which considers a limited number of covariates, high-dimensional matching can involve hundreds of empirical covariates. Another promising approach is leveraging external databases such as ChEMBL. ChEMBL provides valuable information about drug indications, contraindications, and other clinical data. Leveraging external databases like ChEMBL enables researchers to automatically select relevant analytic cohorts and covariates relevant to drug indication and treatment.

## CONCLUSION

We demonstrated the potential of using statistical data mining methods to generate and prioritize hypotheses on medication association with the risk of PTB. This foundational framework can be used for adverse outcomes such as gestational diabetes or preeclampsia. We note that these results should be further validated, reflecting the characteristics of exposures of interest and their indication. We only investigated drug effects associated with the risk of PTB. The mentioned drugs may be attributed to other adverse pregnancy outcomes or congenital disorders.

### Contributors

Yeon Mi Hwang: Conceptualization, Resources, Data Curation, Software, Formal Analysis, Investigation, Visualization, Data Interpretation, Methodology, Writing - original draft, Writing - review and editing; Samantha N. Piekos; Conceptualization, Data Curation, Data Interpretation, Writing - review and editing; Qi Wei: Data curation, Data Interpretation; Nathan D. Price: Conceptualization, Writing - review and editing; Leroy Hood: Conceptualization, Writing - review and editing; Jennifer J Hadlock: Conceptualization, Supervision, Writing - review and editing; YH, QW, SNP, and JJH had full access to the data in the study and verified the data. All authors had final responsibility for the decision to submit for publication.

### Data Sharing Statement

All clinical logic has been shared. Results have been aggregated and reported within this paper to the extent possible while maintaining privacy from personal health information (PHI) as required by law. All data is archived within PSJH systems in a HIPAA-secure audited compute environment to facilitate verification of study conclusions.

## Data Availability

All clinical logic has been shared and available online at https://github.com/Hadlock-Lab/PSM_Maternity_at_scale. Results have been aggregated and reported within this paper to the extent possible while maintaining privacy from personal health information (PHI) as required by law. All data is archived within PSJH systems in a HIPAA-secure audited compute environment to facilitate verification of study conclusions.

## Acknowledgements

We are grateful to the Institute for Systems Biology for startup funds, and to PSJH for sharing their data engineering expertise and computational resources. We would also like to acknowledge SNOMED International for developing and maintaining SNOMED-CT©.

