## Supplementary materials for "Detection of Drug Effect Signals Associated with Adverse Pregnancy Outcomes Using Propensity Score Matching at Scale"

**SUPPLEMENTARY METHODS**

**Method S1** Investigation on the association between exposure to sertraline/SSRI and elevated risk of PTB

We limited our analytic population to patients with at least one depression diagnosis before the pregnancy (Table S2). We calculated the risk of PTB using the propensity score adjustment method. We chose propensity score adjustment because it performs well compared to other covariate adjustment methods[1], and our analytic population shares similar characteristics and is balanced. Propensity score was calculated on maternal and pregnancy characteristics. This includes delivery year, parity, gravidity, maternal age, race, ethnicity, preterm history, ethanol consumption status, smoking status, and illegal drug use status.

**Method S2** Investigation on negative correlation between exposure to acyclovir and PTB risk

We first assessed the risk of PTB in patients exposed to acyclovir before the GA of 36 weeks using propensity score matching. We used matching here because the control and treatment groups were highly imbalanced. Second, we calculated the chance of delivering before 37 weeks on a subsample of patients with an indication of genital herpes, determined based on a diagnosis before delivery (Table S2). Third, we compared PTB risk between patients exposed to acyclovir and valacyclovir before GA 36 weeks. We used the propensity score adjustment method for the second and third analyses because the control and treatment groups were balanced; both the control and treatment groups had genital herpes. Propensity score was calculated on maternal and pregnancy characteristics. This includes delivery year, parity, gravidity, maternal age, race, ethnicity, preterm history, ethanol consumption status, smoking status, and illegal drug use status.

**Method S3** Investigation on the association between exposure to ferrous sulfate and decreased risk of PTB

We limited our analytic population to patients diagnosed with iron deficiency anemia (Table S2) within 180 days of the prepregnancy period.  We used propensity score adjustment because the control and treatment groups were balanced; both had iron deficiency anemia before pregnancy. The propensity score was calculated on maternal/pregnancy characteristics. This includes delivery year, parity, gravidity, maternal age, race, ethnicity, preterm history, ethanol consumption status, smoking status, and illegal drug use status.

**SUPPLEMENTARY TABLES**

**Table S1** Variable definition

| **Category** | **Variable** | **Definitions** |
| --- | --- | --- |
| **Maternal/pregnancy characteristics** | Race | Race noted in medical record. Missing values were encoded as Unknown; American Indian or Alaska Native, Asian, Black or African American, Native Hawaiian or Other Pacific Islander, White or Caucasian, Multiracial, Other, Unknown |
|  | Ethnicity | Ethnicity noted in medical record. Missing values were encoded as unknown; Hispanic or Latino, Not Hispanic or Latino, Unknown |
|  | Maternal age | Maternal age at the start of pregnancy; 18 ~ 24, 25 ~ 29, 30 ~ 34, 35 ~ 40, 41 ~ 44 |
|  | Pregravid BMI | Pregravid Body Mass Index (kg/m2). Missing value was encoded as unknown; Underweight (<18.5 BMI), Normal (18.5 - 24.9 BMI), Overweight ( 25.0 - 29.9 BMI), Obese (>30.0) |
|  | Insurance status | Commercial, Medicaid, Medicare, Uninsured-Self-Pay |
|  | Smoker | Self-reported smoking status; 0,1 |
|  | Illegal drug use | Self-reported illegal drug use status; 0,1 |
|  | Preterm history | History of preterm delivery; 0,1 |
|  | Parity | Number of times a patient has delivered a fetus older than 20 weeks of gestation prior to the current pregnancy; 0, 1~5, 6 |
|  | Gravidity | Number of times a patient has been pregnant; 0, 1~5, 6 |
|  | Delivery year | Year of delivery; 2013~2022 |
| **Maternal-fetal health outcomes** | Low birth weight | Infant birth weight ≤ 2,500g |
|  | Preterm birth | Infant gestational age (GA) at birth < 37 weeks |
|  | Small for gestational age | Infant birth weight < 10th percentiles for infants of same GA |

BMI Body Mass Index; CDC Center for Disease Control and Prevention; GA Gestational age; SVI Social Vulnerability Index

**Table S2** SNOMED diagnosis codes

| **Diagnosis** | **SNOMED Concept ID** |
| --- | --- |
| Abnormal fetal movement | 276369006, 363093002, 364755008 |
| anemia | 271737000 |
| anxiety | 48694002 |
| asthma | 195967001 |
| Bacterial infection | 87628006,442635007,170488007, 710954001 |
| Breech presentation | 6096002 |
| Cardiovascular diseases | 49601007 |
| Chronic kidney diseases | 709044004 |
| Cystic fibrosis | 190905008 |
| Depression | 35489007 |
| Diabetes | 73211009 |
| Excessive fetal growth | 22173004, 199616008 |
| Fatigue | 84229001 |
| Gastroesophageal reflux diseases | 235595009 |
| History of cesarean section | 161805006 |
| Hypertensive disorder | 10725009,720568003,48146000,59621000,429198000, 71701000119105,71421000119105, 397748008, 706882009, 697929007, 1078301000112109, 206596003, 23130000, 5501000119106, 118781000119108, 84094009, 31992008, 38341003, 24184005 69909000, 198941007,  367390009, 48194001 |
| Intrauterine device birth control | 737288007 |
| insomnia | 193462001 |
| Irregular period | 80182007, 237055002 (polycystic ovary syndrome) |
| Leukemia | 93143009 |
| Placenta previa | 36813001, 445122007 |
| Pneumonia | 233604007 |
| Poor fetal growth | 2033007, 397949005 |
| Preterm labor | 289733005, 6383007 |
| Premature rupture of membrane | 44223004 |
| Rhesus D negative | 165746003 |
| sepsis | 91302008 |
| Sickle cell diseases | 417357006 |
| Vitamin D deficiency | 34713006 |

All descendant SNOMED codes are included

**Table S3** Descriptive statistics of source population

|  | **Source population (n=367,459)** |
| --- | --- |
| **Maternal-fetal health outcome** |  |
| **Gestational days** | 275.0 (12.8) |
| **Preterm birth** | 28185 (7.7) |
| **Small for gestational age** | 43905 (12.1) |
| **Low birth weight** | 19611 (5.4) |
| **Delivery method** |  |
| vaginal | 252001 (69.0) |
| c-section | 110386 (30.2) |
| other | 277 (0.1) |
| Missing | 2411 (0.7) |
| **Maternal/pregnancy characteristics** |  |
| **Maternal age** | 31.0 (5.8) |
| **Age group** |  |
| 17 or younger | 3112 (1.1) |
| 18-24 | 61214 (22.1) |
| 25-29 | 93220 (33.6) |
| 30-34 | 119530 (43.1) |
| **Race group** |  |
| American Indian or Alaska Native | 4764 (1.3) |
| Asian | 29448 (8.1) |
| Black or African American | 15478 (4.2) |
| Native Hawaiian or Other Pacific Islander | 4249 (1.2) |
| White or Caucasian | 230765 (63.2) |
| Multirace | 17932 (4.9) |
| Other | 60175 (16.5) |
| Missing | 2264 (0.6) |
| **Ethnic group** |  |
| Hispanic/Latino | 75271 (20.6) |
| Not Hispanic/Latino | 281675 (77.2) |
| Unknown/Not Reported | 8061 (2.2) |
| **BMI category** |  |
| underweight | 3002 (0.8) |
| normal | 52389 (14.4) |
| obese | 31243 (8.6) |
| overweight | 31300 (8.6) |
| Missing | 247141 (67.7) |
| **Insurance** |  |
| Commercial | 170164 (46.6) |
| Medicaid/Medicare | 192453 (52.7) |
| Self Pay | 84 (0.0) |
| Missing | 2374 (0.7) |
| **Smoker** | 26108 (7.2) |
| **Illegal drug user** | 27071 (7.4) |
| **Alcohol user** | 64577 (17.7) |
| **Vulnerability index of socioeconomic status** | 0.4 (0.2) |
| **Vulnerability index of housing composition** | 0.4 (0.3) |
| **Vulnerability index of minority status and language** | 0.6 (0.2) |
| **Vulnerability index of housing type and transportation** | 0.6 (0.3) |
| **Rural/urban classification** |  |
| Metropolitan | 307464 (84.2) |
| Micropolitan | 17425 (4.8) |
| Small Town | 5871 (1.6) |
| Rural | 4567 (1.3) |
| Missing | 29747 (8.1) |
| **Gravidity** |  |
| 1 | 82176 (22.5) |
| 2-4 | 198452 (54.4) |
| 5≤ | 42011 (11.5) |
| Missing | 42436 (11.6) |
| **Parity** |  |
| 0 | 45275 (12.4) |
| 1 | 127735 (35.0) |
| 2-4 | 141207 (38.7) |
| 5≤ | 8422 (2.3) |
| Missing | 42436 (11.6) |
| Preterm history |  |
| Yes | 76632 (21.0) |
| No | 246007 (67.4) |
| Missing | 42436 (11.6) |
| **Fetal sex** |  |
| Female | 164049 (48.6) |
| Male | 173160 (51.3) |
| Unknown | 51 (0.0) |
| Other | 16 (0.0) |
| **Delivery year** |  |
| 2013 | 20799 (5.9) |
| 2014 | 30834 (8.8) |
| 2015 | 34844 (10.0) |
| 2016 | 36288 (10.4) |
| 2017 | 35895 (10.3) |
| 2018 | 35587 (10.2) |
| 2019 | 36953 (10.6) |
| 2020 | 35431 (10.1) |
| 2021 | 39791 (11.4) |
| 2022 | 43188 (12.4) |

**Table S4** Categorization of medications with statistically significant association with risk of PTB based on their indication

| **Medication** | **Usage/usage during pregnancy** | **Indication** | | |
| --- | --- | --- | --- | --- |
|  |  | **Preterm labor/birth** | **PTB risk factor** | **Infection** |
| Ursodeoxyholate | Dissolve gallstones. Treat cholestatic liver diseases[2] | N | Y(liver disorder[3]) | N |
| Indomethacin | Pain treatment. Tocolytics for preterm labor[4] | Y[4] | N | N |
| Enoxaparin | Anticoagulation[5] | N | Y(blood clot[6]) | N |
| Progesterone | Treatment option for preterm birth[7] | Y | N | N |
| Diazepam | Treat anxiety, muscle spasm, and seizures[8] | N | Y(anxiety[9]) | N |
| Insulin isophane beef | Insulin given to help control blood sugar levels in people with diabetes[10] | N | Y(diabetes[11]) | N |
| Diclofenac | Treat pain, migraines, and arthritis[12] | N | Y (arthritis[13]) | N |
| Sucralfate | Medication used to treat duodenal ulcers, chemotherapy-induced mucostasis | N | Y(cancer[14]) | N |
| Nifedipine | Treat high blood pressure[15] | Y[16] | Y (diabete[11]) | N |
| Potassium chloride | Treat and prevent low blood potassium[17] | N | Y(hypokalemia[18]) | N |
| Insulin aspart human | Treat diabetes[19] | N | Y(diabetes[11]) | N |
| Sulfamethoxazole | Antibiotics used for bacterial infection such as urinary tract infection[20] | N | Y(urinary tract infection[21]) | Y |
| Clonazepam | Sedative. Treatment for seizures, panic disorder, and anxiety[22] | N | Y(anxiety[23], panic[24]) | N |
| Methylprednisolone | Treatment for inflammation, flares of chronic illness[25] | Y[26] | N | N |
| Prochlorperazine | Treatment for nausea, vomiting, anxiety, and schizophrenia[27] | N | Y(schizophrenia[28]) | N |
| Doxycycline | Antibiotics used for bacterial infection including acne, urinary tract infection, gonorrhea, chlamydia, etc[29] | N | Y(chlamydia[30]) | Y |
| Phenazopyridine | Relieve symptoms caused by urinary tract infection[31] | N | Y(urinary tract infection[21]) | Y |
| Ascorbic acid | Treat vitamin c deficiency[32] | N | Y[33] | N |
| Epinephrine | Treatment for severe asthma attack and allergic reactions[34] | N | Y(asthma[35] | N |
| Insulin lispro | Treament for diabetes[36] | N | Y[11] | N |
| Nystatin | Treat fungal infection[37] | N | Y(candida[38]) | Y |
| Clindamycin | Antibiotic for skin and vaginal infection[39] | N | Y[40] | Y |
| Ferrous gluconate | Used to treat or prevent iron deficiency anemia[41] | N | Y(anemia[42]) | N |
| Pantoprazole | Reduce the amount of stomach acid. Used for heartburn, acid reflux, gastroesophegal reflux diseases[43] | N | Y(gerd[44]) | N |
| Trazodone | Antidepresssant[45] | N | Y(depression[46]) | N |
| Tramadol | Strong pain medication[47] | N | N | N |
| Benzonatate | Treat cough caused by common cold and other breathing problems(asthma, pneumonia)[48] | N | Y[35] | N |
| Levothyroxine | Treat hypothyroidism[49] | N | Y(hypothyroidism[50]) | N |
| Metochloropramide | Treat gerd, and gastroparesis in patients with diabetes[51] | N | Y[44] | N |
| Metformin | Treat type 2 diabetes[52] | N | Y(diabetes[11]) | N |
| Sertraline | Antidepressant, selective serotonin reuptake inhibitor[53] | N | Y(depression[9]) | N |
| Promethazine | Antihistamine treating allergies and motion sickness[54] | N | N | N |
| Polyethylene glycol 3350 | Treat occasional constipation[55] | N | N | N |
| Azithromycin | Antibiotic medicine treating pneumonia, ear, nose, throat, and sinus[56] | N | Y(pneumonia[57]) | Y |
| Acetaminophen | Treat minor aches, pains, and fevers[58] | N | N | N |
| Famotidine | Used to treat gerd[59] | N | Y(gerd[44]) | N |
| Amoxicillin | Penicillin antibiotic treating infection and stomach ulcers[60] | N | N | Y |
| Norethindrone | Used to prevent pregnancy[61] | N | N | N |
| Insulin isophane | Control blood sugar with diabetes[10] | N | Y(diabetes[11]) | N |
| Oxycodone | Treat moderate to severe pain[62] | N | N | N |
| Ondansetron | Prevent nausea and vomiting[63] | N | N | N |
| Ibuprofen | Treat fever and mild to severe pain[64] | N | N | N |
| Hydrocortisone | Calm down body's immune response to reduce pain, itching, swelling, inflammation[65] | N | N | N |
| Ferrous sulfate | Iron supplement used for iron deficient anemia[66] | N | Y[42] | N |
| Clavulanate | Used in conjuction with amoxicillin to treat certain bacterial infection[67] | N | N | Y |
| Pseudoephedrine | Treat stuffy nose and sinuses[68] | N | N | N |
| Glyburide | Treat type 2 diabetes[69] | N | Y[11] | N |
| Vitamin b6 | Vitamin for healthy nervous system and immune system[70] | N | N | N |
| Tretinoin | Acne treatment[71] | N | N | N |
| Glucose oxidase | Probe for sensing gestational diabetes[72] | N | N | N |
| Acyclovir | Treat herpes virus infection[73] | N | Y(genital herpes[74]) | Y |
| Etonogestrel | Birth control[75] | N | N | N |
| Pramoxine | Relieve pain and itching from insect bites or hemorrhoids[76] | N | N | N |

**SUPPLEMENTARY FIGURES**

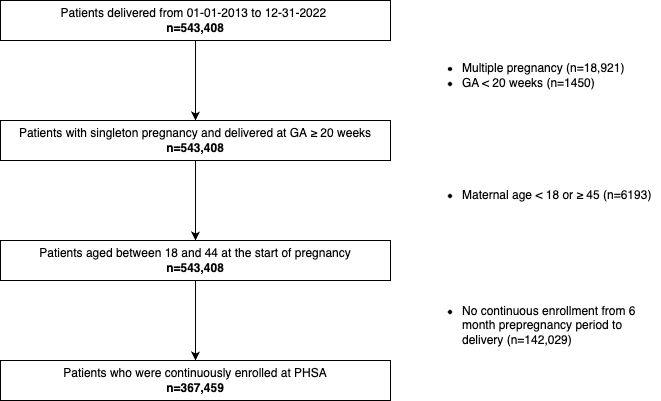
**Figure S1** Source population selection flow diagram

**Figure S2** Maternal age distribution from 2013 to 2022

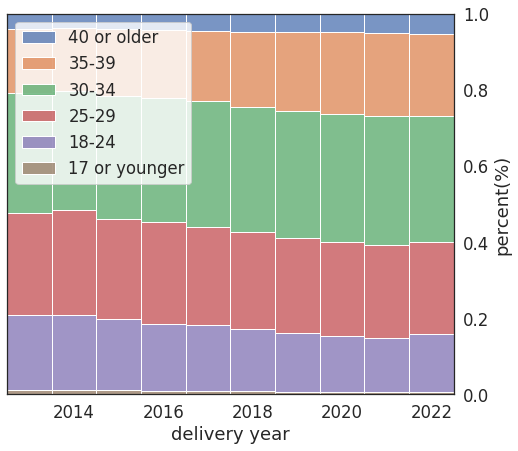

**Figure S3** Prescription rate of medication based on RxNorm ingredient and ATC category

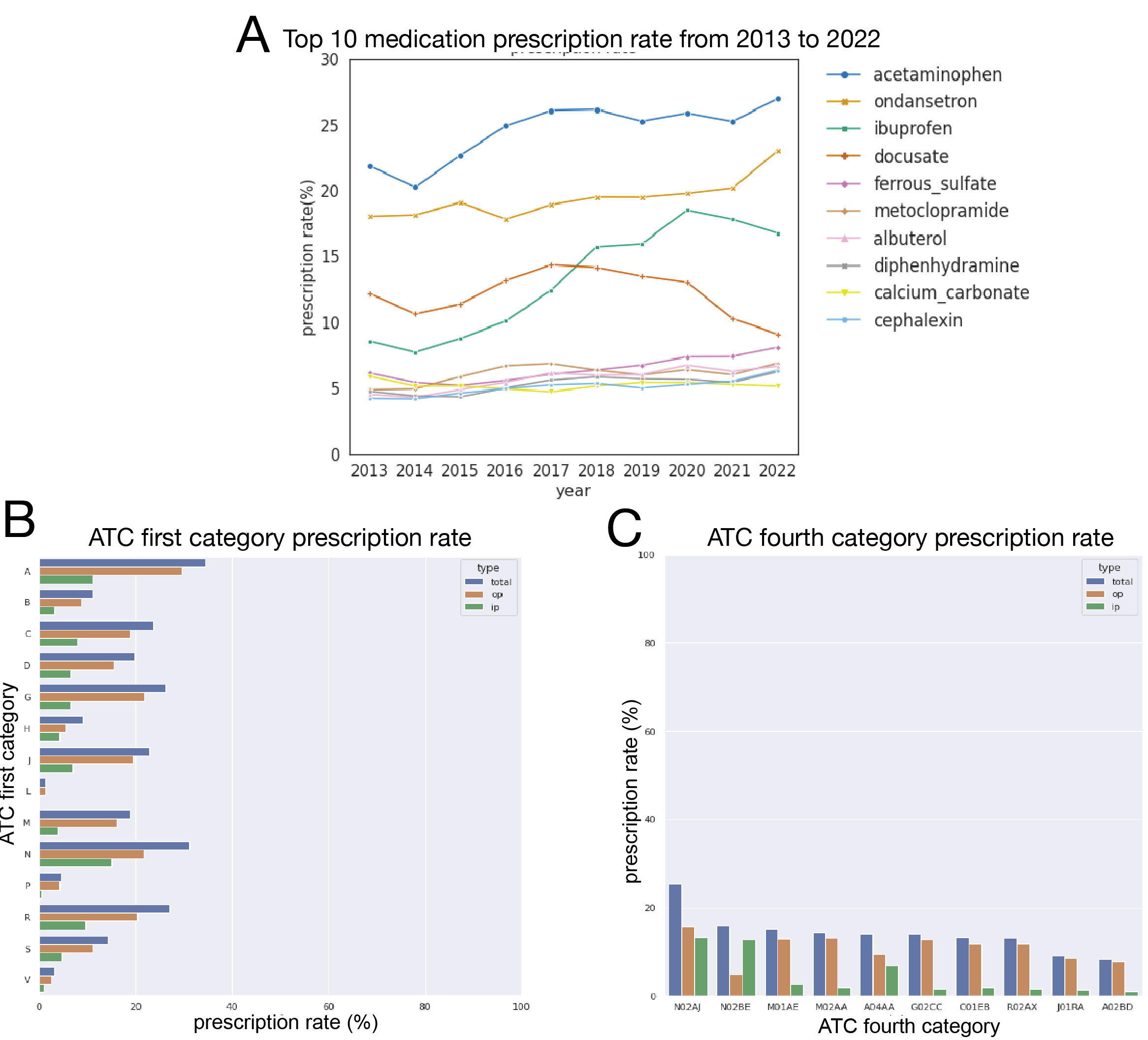

A. Prescription rate of top ten commonly prescribed medications from 2013 to 2022. Top ten commonly prescribed medications were acetaminophen, ondansetron, ibuprofen, docusate, ferrous sulfate, metoclopramide, albuterol, and diphenhydramine in order.
A. Prescription rate of top ten commonly prescribed medication based on ATC first level. ATC first level has fourteen main anatomical or pharmacological groups. A: Alimnetary tract and metabolism; B: Blood and blood forming organs; C: Cardiovascular system; D: Dermatologicals; G: Genito urinary system and sex hormones; H: Systemic hormonal preparations, excluding sex hormones and insulins; J: Antiinfective for systemic use; L: Antineoplastic and immunomodulating agents; M: Musculoskeletal system; N: Nervous system; P: Antiparasitic products, insecticides and repellants; R: Respiratory system; S: Sensory organs; V: Various.[77]
A. Prescription rate of top ten commonly prescribed medication based on ATC fourth level. ATC fourth level indicates chemical subgroup. N02AJ: Opioids in combination with non-opiod analgesics; N02BE: Anilides; M01AE: Propionics acid derivative; M02AA: Antiinflammatory preparations, non-steroids for topical use; A04AA: Serotonin antagonist; G02CC: Antiinflammatory products for vaginal administration; C01EB: Other cardiac preparation; R02AX: Other throat preparation; J01RA: Combinations of antibacterials; A02BD: Combinations for eradications of helicobacter pylori

1	Elze MC, Gregson J, Baber U, *et al.* Comparison of Propensity Score Methods and Covariate Adjustment: Evaluation in 4 Cardiovascular Studies. *J Am Coll Cardiol* 2017;**69**:345–57.

2	Luketic VA, Sanyal AJ. The current status of ursodeoxycholate in the treatment of chronic cholestatic liver disease. *Gastroenterologist* 1994;**2**:74–9.

3	Mawson AR. A Role for the Liver in Parturition and Preterm Birth. *J Transl Sci* 2016;**2**:154–9.

4	Haas DM, Benjamin T, Sawyer R, *et al.* Short-term tocolytics for preterm delivery - current perspectives. *Int J Womens Health* 2014;**6**:343–9.

5	Jupalli A, Iqbal AM. *Enoxaparin*. StatPearls Publishing 2022.

6	Flickr F us on. What are the risk factors for preterm labor and birth? https://www.nichd.nih.gov/. https://www.nichd.nih.gov/health/topics/preterm/conditioninfo/who_risk (accessed 13 Sep 2023).

7	Updated Clinical Guidance for the Use of Progesterone Supplementation for the Prevention of Recurrent Preterm Birth. https://www.acog.org/clinical/clinical-guidance/practice-advisory/articles/2023/04/updated-guidance-use-of-progesterone-supplementation-for-prevention-of-recurrent-preterm-birth (accessed 13 Sep 2023).

8	Dhaliwal JS, Rosani A, Saadabadi A. *Diazepam*. StatPearls Publishing 2022.

9	Männistö T, Mendola P, Kiely M, *et al.* Maternal psychiatric disorders and risk of preterm birth. *Ann Epidemiol* 2016;**26**:14–20.

10	Saleem F, Sharma A. *NPH Insulin*. StatPearls Publishing 2023.

11	Crump C, Sundquist J, Sundquist K. Preterm birth and risk of type 1 and type 2 diabetes: a national cohort study. *Diabetologia* 2020;**63**:508–18.

12	Alfaro RA, Davis DD. *Diclofenac*. StatPearls Publishing 2023.

13	Smith CJF, Förger F, Bandoli G, *et al.* Factors Associated With Preterm Delivery Among Women With Rheumatoid Arthritis and Women With Juvenile Idiopathic Arthritis. *Arthritis Care Res* 2019;**71**:1019–27.

14	Van Calsteren K, Heyns L, De Smet F, *et al.* Cancer during pregnancy: an analysis of 215 patients emphasizing the obstetrical and the neonatal outcomes. *J Clin Oncol* 2010;**28**:683–9.

15	Khan KM, Patel JB, Schaefer TJ. *Nifedipine*. StatPearls Publishing 2023.

16	Conde-Agudelo A, Romero R, Kusanovic JP. Nifedipine in the management of preterm labor: a systematic review and metaanalysis. *Am J Obstet Gynecol* 2011;**204**:134.e1-134.e20.

17	McMahon RS, Bashir K. *Potassium Chloride*. StatPearls Publishing 2023.

18	Yang C-W, Li S, Dong Y. The Prevalence and Risk Factors of Hypokalemia in Pregnancy-Related Hospitalizations: A Nationwide Population Study. *Int J Nephrol* 2021;**2021**:9922245.

19	Rubin R, Khanna NR, McIver LA. *Aspart Insulin*. StatPearls Publishing 2022.

20	Kemnic TR, Coleman M. *Trimethoprim Sulfamethoxazole*. StatPearls Publishing 2022.

21	Baer RJ, Nidey N, Bandoli G, *et al.* Risk of Early Birth among Women with a Urinary Tract Infection: A Retrospective Cohort Study. *AJP Rep* 2021;**11**:e5–14.

22	Basit H, Kahwaji CI. *Clonazepam*. StatPearls Publishing 2023.

23	Rose MS, Pana G, Premji S. Prenatal Maternal Anxiety as a Risk Factor for Preterm Birth and the Effects of Heterogeneity on This Relationship: A Systematic Review and Meta-Analysis. *Biomed Res Int* 2016;**2016**:8312158.

24	Yonkers KA, Gilstad-Hayden K, Forray A, *et al.* Association of Panic Disorder, Generalized Anxiety Disorder, and Benzodiazepine Treatment During Pregnancy With Risk of Adverse Birth Outcomes. *JAMA Psychiatry* 2017;**74**:1145–52.

25	Ocejo A, Correa R. *Methylprednisolone*. StatPearls Publishing 2022.

26	Crowley P. Prophylactic corticosteroids for preterm birth. *Cochrane Database Syst Rev* 2000;:CD000065.

27	Din L, Preuss CV. *Prochlorperazine*. StatPearls Publishing 2023.

28	Fabre C, Pauly V, Baumstarck K, *et al.* Pregnancy, delivery and neonatal complications in women with schizophrenia: a national population-based cohort study. *Lancet Reg Health Eur* 2021;**10**:100209.

29	Patel RS, Parmar M. *Doxycycline Hyclate*. StatPearls Publishing 2023.

30	Rours GIJG, Duijts L, Moll HA, *et al.* Chlamydia trachomatis infection during pregnancy associated with preterm delivery: a population-based prospective cohort study. *Eur J Epidemiol* 2011;**26**:493–502.

31	Eastham JH, Patel P. *Phenazopyridine*. StatPearls Publishing 2023.

32	Maxfield L, Crane JS. *Vitamin C Deficiency*. StatPearls Publishing 2022.

33	Hauth JC, Clifton RG, Roberts JM, *et al.* Vitamin C and E supplementation to prevent spontaneous preterm birth: a randomized controlled trial. *Obstet Gynecol* 2010;**116**:653–8.

34	Dalal R, Grujic D. *Epinephrine*. StatPearls Publishing 2023.

35	Kelly YJ, Brabin BJ, Milligan P, *et al.* Maternal asthma, premature birth, and the risk of respiratory morbidity in schoolchildren in Merseyside. *Thorax* 1995;**50**:525–30.

36	Islam N, Khanna NR, Zito PM. *Insulin Lispro*. StatPearls Publishing 2023.

37	*Nystatin*. National Institute of Diabetes and Digestive and Kidney Diseases 2020.

38	Maki Y, Fujisaki M, Sato Y, *et al.* Candida Chorioamnionitis Leads to Preterm Birth and Adverse Fetal-Neonatal Outcome. *Infect Dis Obstet Gynecol* 2017;**2017**:9060138.

39	Murphy PB, Bistas KG, Le JK. *Clindamycin*. StatPearls Publishing 2023.

40	McDonald HM, O’Loughlin JA, Jolley P, *et al.* Vaginal infection and preterm labour. *Br J Obstet Gynaecol* 1991;**98**:427–35.

41	Ferrous gluconate. Drugs.com. https://www.drugs.com/mtm/ferrous-gluconate.html (accessed 13 Sep 2023).

42	Rahmati S, Azami M, Badfar G, *et al.* The relationship between maternal anemia during pregnancy with preterm birth: a systematic review and meta-analysis. *J Matern Fetal Neonatal Med* 2020;**33**:2679–89.

43	Bernshteyn MA, Masood U. *Pantoprazole*. StatPearls Publishing 2023.

44	Lee K-S, Kim ES, Kim D-Y, *et al.* Association of Gastroesophageal Reflux Disease with Preterm Birth: Machine Learning Analysis. *J Korean Med Sci* 2021;**36**:e282.

45	Shin JJ, Saadabadi A. *Trazodone*. StatPearls Publishing 2022.

46	Grote NK, Bridge JA, Gavin AR, *et al.* A meta-analysis of depression during pregnancy and the risk of preterm birth, low birth weight, and intrauterine growth restriction. *Arch Gen Psychiatry* 2010;**67**:1012–24.

47	Dhesi M, Maldonado KA, Maani CV. *Tramadol*. StatPearls Publishing 2023.

48	Philip Thornton D. Benzonatate. Drugs.com. https://www.drugs.com/benzonatate.html (accessed 13 Sep 2023).

49	Eghtedari B, Correa R. *Levothyroxine*. StatPearls Publishing 2022.

50	Parizad Nasirkandy M, Badfar G, Shohani M, *et al.* The relation of maternal hypothyroidism and hypothyroxinemia during pregnancy on preterm birth: An updated systematic review and meta-analysis. *Int J Reprod Biomed* 2017;**15**:543–52.

51	Isola S, Hussain A, Dua A, *et al.* *Metoclopramide*. StatPearls Publishing 2023.

52	Nasri H, Rafieian-Kopaei M. Metformin: Current knowledge. *J Res Med Sci* 2014;**19**:658–64.

53	Singh HK, Saadabadi A. *Sertraline*. StatPearls Publishing 2023.

54	Southard BT, Al Khalili Y. *Promethazine*. StatPearls Publishing 2022.

55	Dabaja A, Dabaja A, Abbas M. *Polyethylene Glycol*. StatPearls Publishing 2023.

56	Sandman Z, Iqbal OA. *Azithromycin*. StatPearls Publishing 2023.

57	Lim WS, Macfarlane JT, Colthorpe CL. Pneumonia and pregnancy. *Thorax* 2001;**56**:398–405.

58	Gerriets V, Anderson J, Nappe TM. *Acetaminophen*. StatPearls Publishing 2023.

59	Nguyen K, Dersnah GD, Ahlawat R. *Famotidine*. StatPearls Publishing 2022.

60	Akhavan BJ, Khanna NR, Vijhani P. *Amoxicillin*. StatPearls Publishing 2022.

61	Norethindrone. Drugs.com. https://www.drugs.com/mtm/norethindrone.html (accessed 13 Sep 2023).

62	Sadiq NM, Dice TJ, Mead T. *Oxycodone*. StatPearls Publishing 2022.

63	Griddine A, Bush JS. *Ondansetron*. StatPearls Publishing 2023.

64	Ngo VTH, Bajaj T. *Ibuprofen*. StatPearls Publishing 2023.

65	Hydrocortisone. Drugs.com. https://www.drugs.com/mtm/hydrocortisone.html (accessed 13 Sep 2023).

66	Sinha S. Ferrous Sulfate: Uses, Dosage & Side Effects. Drugs.com. https://www.drugs.com/ferrous_sulfate.html (accessed 13 Sep 2023).

67	Uto LR, Gerriets V. *Clavulanic Acid*. StatPearls Publishing 2023.

68	Pseudoephedrine: medicine for a stuffy or blocked nose. nhs.uk. https://www.nhs.uk/medicines/pseudoephedrine/ (accessed 13 Sep 2023).

69	Hardin MD, Jacobs TF. *Glyburide*. StatPearls Publishing 2023.

70	Vitamin B6. https://ods.od.nih.gov/factsheets/VitaminB6-HealthProfessional/ (accessed 13 Sep 2023).

71	Schmidt N, Gans EH. Tretinoin: A Review of Its Anti-inflammatory Properties in the Treatment of Acne. *J Clin Aesthet Dermatol* 2011;**4**:22–9.

72	Liu B, Dai Q, Liu P, *et al.* Nanostructure-mediated glucose oxidase biofunctionalization for monitoring gestational diabetes. *Process Biochem* 2021;**110**:19–25.

73	Taylor M, Gerriets V. *Acyclovir*. StatPearls Publishing 2023.

74	Li D-K, Raebel MA, Cheetham TC, *et al.* Genital herpes and its treatment in relation to preterm delivery. *Am J Epidemiol* 2014;**180**:1109–17.

75	Gilbert AL, Hoffman BL. Contraceptive Technology: Present and Future. *Obstet Gynecol Clin North Am* 2021;**48**:723–35.

76	Pramoxine. Memorial Sloan Kettering Cancer Center. https://www.mskcc.org/cancer-care/patient-education/medications/adult/pramoxine (accessed 13 Sep 2023).

77	Anatomical Therapeutic Chemical (ATC) Classification. https://www.who.int/tools/atc-ddd-toolkit/atc-classification (accessed 21 Nov 2021).
